# Development and clinical evaluation of a monkeypox antigen-detecting rapid diagnostic test

**DOI:** 10.1101/2024.10.06.24313376

**Authors:** Nobuyuki Kurosawa, Tatsuhiko Ozawa, Kousei Ozawa, Masayuki Shimojima, Madoka Kawahara, Fumi Kasuya, Wakaba Okada, Mami Nagashima, Kenji Sadamasu, Masae Itamochi, Hideki Tani, Yoshitomo Morinaga, Kosuke Yuhara, Jun Okamoto, Haruna Ichikawa, Takashi Kawahata, Tomomi Yamazaki, Masaharu Isobe

## Abstract

To address the global emergence of monkeypox after the 2022 epidemic, a rapid and accurate diagnostic tool is needed at the point of care to identify individuals infected with monkeypox virus (MPXV) to prevent and control the spread of the virus. We designed an antigen-detecting rapid diagnostic test that exclusively detects MPXV without cross-reacting with the vaccinia virus by developing monoclonal antibodies against the MPXV nuclear capsid protein A5L (MPXV-A5L). The test established a limit of detection sensitivity of 0.5 ng/mL of MPXV-A5L, with high sensitivity (87%) for clinical specimens collected from MPXV patients, a qPCR cycle threshold value ≤ 25 and 100% specificity for qPCR-negative samples. The test is an ideal rapid diagnostic tool for supporting clinical decision-making for people suspected of having MPXV infection in resource-poor settings.

## 1. Introduction

Monkeypox is a zoonotic disease caused by the monkeypox virus (MPXV), a highly pathogenic double-stranded DNA virus classified in the genus orthopoxvirus (OPXV) of the family *Poxviridae* (Karagoz et al., 2023; Mitja et al., 2023; Yu, Shi, and Cheng, 2023). There are two genotypes of MPXV: Congo Basin strains (clade I) and West African strains (clades IIa and IIb). Although monkeypox was confirmed mainly in African countries, an epidemic of clade IIb MPXV was ongoing globally in early 2022, and 88,288 infected people and approximately 150 deaths were reported from 112 countries on July 10, 2023 (Malik et al., 2023). Infection occurs through close contact with respiratory droplets, body fluids, skin lesions, or the blood of infected people or animals. After an incubation period ranging from 7 to 16 days, the affected individuals experience prodromal symptoms such as fever and general malaise, followed by the appearance of a skin rash that can be mistaken for those seen in chickenpox, measles, syphilis, and hand-foot-and-mouth disease (Zahmatyar et al., 2023) (Khattak et al., 2022) (Silva et al., 2023). Most healthy individuals experience mild disease progression, but immunocompromised patients, children, and pregnant women can experience severe symptoms (Schwartz et al., 2023; Silva et al., 2023). Older adults who have received smallpox vaccination acquired cross-protective immunity against MPXV. However, younger generations, who had not been vaccinated against smallpox, have no immunity to MPXV (Reynolds and Damon, 2012). Although the WHO declared the end of the monkeypox emergency, it called for sustained efforts for the long-term management of this virus (Bunge et al., 2022).

The detection of viral genes via polymerase chain reaction (PCR) is the gold standard method for MPXV diagnosis (Huang et al., 2023). However, these methods are designed for use in centralized laboratory settings. An antigen-detecting rapid diagnostic test (Ag-RDT) based on a lateral flow immunochromatography assay (LFIA) is an effective method for diagnosing MPXV through point-of-care testing in patients with monkeypox symptoms (Bunge et al., 2022). Several commercial and in-house Ag-RDTs have been developed for the diagnosis of MPXV by using antibodies against vaccinia virus (VACV) A27 or MPXV A29, an ortholog of VACV A29 (Mitja et al., 2023) (Huang et al., 2023; Hughes et al., 2014; Roumillat, Patton, and Davis, 1984). However, it is difficult to diagnose MPXV accurately, as most tests are not specific and may cross-react with VACV, which circulates in wild and domestic animals with occasional transmission to humans in close contact with these animals. Thus, an accurate MPXV Ag-RDT is vital for the effective treatment and control of MPXV infection, especially in areas where VACV is endemic (Miranda et al., 2017).

MPXV A5L, an ortholog of VACV A4L, is a highly conserved 39-kDa immunodominant nucleocapsid protein that plays a role in the assembly and disassembly of the virion (Risco et al., 1999). The abundance and low mutation rate of MPXV A5L make it an ideal antigen for detecting MPXV via LFIA. In this work, we designed an MPXV Ag-RDT by developing a set of monoclonal antibodies that target MPXV A5L while not cross-reacting with VACV. We confirmed assay specificity via a panel of viral and bacterial pathogens. The MPXV Ag-RDT showed high sensitivity (87%) for clinical specimens collected from MPXV patients, with a qPCR cycle threshold (Ct) value ≤ 25 and a specificity of 100% for qPCR-negative samples. The MPXV Ag-RDT is suitable for point-of-care use in low-resource settings and can support the implementation of isolation measures to reduce the risk of transmission.

## 2. Materials and methods

### 2.1 Ethics approval

All animal and biosafety experiments were carried out at the University of Toyama and Toyama Institute of Health after approval from the Committee for Laboratory Animal Care and Use (A2022ENG-1 and G2022eng-1). Experiments with live VACV and MPXV were performed under biosafety levels 2 and 3, respectively, at the National Institute of Infectious Diseases. The clinical study was approved by the Ethical Review Board of the Tokyo Metropolitan Institute of Public Health (5 KenKenKen 3712).

### 2.2 Antigen preparation

Chemically synthesized MPXV A5L and VACA A4L cDNA were inserted into the pCold ProS2 vector (Takara Bio, Japan) and transformed into *E. coli* Tuner (DE3) pLacI (Novagen). Recombinant proteins were purified from the soluble fraction of the *E. coli* lysate via Ni-chelate affinity chromatography (Capturem His-tagged Purification Miniprep Kit, Takara Bio, Japan) followed by dialysis with PBS. The purified proteins were then concentrated using Amicon® Ultra-15 centrifugal filter units (EMD Millipore) and used as antigens. The MPXV peptide (LKDLMSSVEKDMRQLQAET) was conjugated to keyhole limpet hemocyanin (KLH) via the C-terminal cysteine residue.

### 2.3 Immunization

Anesthetized rats or rabbits were immunized three times intramuscularly at the tail base with 50 μg of A5L protein. Iliac lymph nodes were surgically removed from the euthanized animals. Anesthetized New Zealand White rabbits were immunized three times subcutaneously with 500 μg of KLH conjugates of MPXV A5L peptide, and peripheral blood lymphocytes were surgically corrected from the euthanized animals.

### 2.4 Development of monoclonal antibodies

The isolation of rat antigen-specific plasma cells was performed as previously described (Kurosawa et al., 2012). Briefly, rat lymph node cells fixed with ice-cold phosphate-buffered saline (PBS) containing 2% paraformaldehyde were stained with DyLight 488-labeled ProS-MPXV-A5L, DyLight 550-labeled ProS-Vaccinia-A4L, DyLight 650-labeled anti-rat IgG and DAPI in PBS containing 0.1% Triton X-100 (PBST). MPXV A5L-specific plasma cells, defined as ProS-MPXV-A5L^High^ and IgG^High^, were single-sorted via a JSAN Cell Sorter (Bay Bioscience). The isolation of rabbit antigen-specific plasma cells was performed as previously described (Ozawa et al., 2012). Briefly, IgG^+^ cells isolated from A5L-immunized rabbits were cultured on a chip coated with 10 μg/mL rabbit IgG antibody at 37 °C for 3 hours. The chip was incubated with biotinylated MPXV A5L peptide (10 μg/mL) for 30 min and stained with Cy3-labeled streptavidin to detect antigen-specific plasma cells. Molecular cloning of immunoglobulin heavy and light chain genes from single-isolated plasma cells, followed by the expression of recombinant antibodies, was performed as previously described (Kurosawa et al., 2012; Ozawa et al., 2012). Antibodies were expressed via the Expi293 Expression System (Thermo Fisher Scientific) and purified using protein A affinity chromatography followed by size-exclusion chromatography (Cytiva).

### 2.5 Immunofluorescence analysis

A plasmid encoding Vaccinia A4L and a series of MPXV A5L mutants were constructed via PCR and inserted into the pCDNA3.1 or pEFMycHis plasmid. Each plasmid was transfected into HEK293 cells using the FuGENE 6 Transfection Reagent (Promega), and the cells were cultured for two days. After fixation with PBS containing 4% paraformaldehyde, the cells were permeabilized with PBST and stained with the indicated antibodies. Images were captured with an Operetta High Content Imaging System (PerkinElmer) or a BZ-X700 all-in-one fluorescence microscope (Keyence).

### 2.6 Preparation of a lateral flow immunochromatographic assay

The #230-9 antibody (0.1 mg) and 100 μL of NanoAct cellulose particles A (BL2: Dark Navy, Asahi Kasei Corporation) were mixed in 900 μL of 10 mM Tris-HCl buffer (pH 8.0) and allowed to stand at 37 °C for 120 minutes. The reaction was stopped by the addition of 12 mL of 100 mM boric acid buffer (pH 8.0) containing 1% casein. The antibody-cellulose particles suspended with 8 mL of 62 mM boric acid buffer (pH 9.2) containing 15% sucrose were uniformly applied to the entire surface of a glass fiber diagnostic pad (GFDX001050, Millipore) at an application rate of 60 μL/cm, followed by drying at 45 °C for 30 minutes to prepare the conjugation pad. #2S-36 and goat-derived anti-rabbit IgG polyclonal antibodies were applied to Hi-Flow Plus 120 Membrane Cards (HF120, Millipore) at predetermined locations in a volume of 1.0 μL/cm with a line width of approximately 1 mm. The membrane was dried for 30 min at 40 °C. The conjugate pad was overlaid onto the nitrocellulose membrane with a 3 mm overlap. The sample pad and the absorbent pad were placed on either end of the nitrocellulose membrane with 3 mm and 5 mm of overlap, respectively. The immunochromatographic specimens were cut into strips 4 mm wide and 60 mm long and placed in a plastic cassette (K007, Shengfeng Plastic).

### 2.7 Immunochromatographic device sensitivity evaluation

Recombinant MPXV A5L was used as a quality control throughout the test manufacturing process. Recombinant MPXV A5L and virus-infected cell lysate were diluted with sample diluent solution (100 mM Tris buffer, pH 8.5, 0.877% sodium chloride, 0.2% Tween 20 and 0.9% Triton X), and an aliquot of 110 μL was dispensed via pipette into the sample well of the cassette. After 15 minutes, the presence of lines was visually checked. Immediately after visual inspection, images were captured, and the signal intensity at both the test and control line locations was quantified using ImageJ (NIH).

### 2.8 Viruses

VACV and MPXV were propagated and titrated as described previously with a modification in that RK13 cells were used instead of Vero cells (Saijo et al., 2006). The SPL-2A7 strain (clade IIb) of MPXV was isolated using RK13 cells from specimens derived from patients with suspected monkeypox and brought to the National Institute of Infectious Diseases for an administrative inspection of monkeypox.

### 2.9 ELISA

RK13 cells infected with VACV or MPXV were washed with PBS and treated with 1% NP40-containing PBS for cell lysis. Uninfected cells were also prepared and treated in parallel. The supernatants of the cell lysates were used as antigens for ELISA. ELISA plates (Nunc Maxisorp) were coated with cell lysates at a 1:1,000 dilution overnight, and blocking was performed with 5% skim milk. The plates were incubated with primary antibodies at the indicated concentrations, followed by further incubation with secondary antibodies labeled with horseradish peroxidase. Colorization was performed by the addition of ABTS (Roche), and the absorbance was measured at 405 nm with a microplate reader (Bio-Rad iMark).

### 2.10 Clinical sample collection and real-time qPCR assay

The samples sent to the Tokyo Metropolitan Institute of Public Health (TMIPH) as a part of the active epidemiological surveillance of suspected cases of monkeypox were used for this study. Viral DNA was extracted from the samples (skin lesions: crusts and blister swabs) via a QIAamp DNA Mini Kit (Qiagen, Hilden, Germany). Real-time qPCR to detect MPXV (targeting the F3L region) was performed using a QuantStudio 12K Flex (Thermo Fisher Scientific, Waltham, MA, USA) and a QuantiTect Probe PCR Kit (Qiagen) according to the pathogen detection manual of the National Institute of Infectious Diseases, Japan (https://www.niid.go.jp/niid/images/lab-manual/mpox20230531.pdf). The viral load of the MPXV was expressed as the Ct value detected by real-time PCR as previously described (Kasuya et al., 2023).

### 2.11 Statistical analysis

The sensitivity, specificity, positive predictive value, and negative predictive value were estimated via the Clopper□Pearson exact method, along with corresponding 95% confidence intervals.

## 3. Results

### 3.1 Development and screening of mAbs against A5L

To develop anti-MPXV A5L antibodies, we immunized rats with recombinant MPXV A5L and rabbits with the KLH-conjugated MPXV A5L peptide. Immunoglobulin heavy and light chain gene cloning from antigen-specific single plasma cells resulted in the production of 128 rat and three rabbit mAbs. The activity of these mAbs was examined via immunofluorescence staining of HEK293 cells expressing MPXV A5L. Among the 61 MPXV A5L-positive mAbs tested, five rat mAb clones and one rabbit mAb clone were selected on the basis of their unique complementarity-determining region 3 sequences. Epitope mapping of these mAbs by using HEK293 cells expressing a series of N-terminal deletion mutants of MPXV A5L revealed that the epitope for #2-22 was located on amino acids 1-100, #2S-36 on amino acids 100-180, #1-28 on amino acids 180-225 and #1-35, #3-7, and #230-9 on amino acids 225-281 (Fig. 1). For further characterization of these mAbs, an ELISA screen was performed using MPXV- or VACV-infected Vero cell lysates as antigens (Fig. 1B). #1-28, #1-35, #2-22, #3-7, and #230-9 reacted with the MPXV- and VACV-infected cell lysates. #2S-36 reacted with only the MPXV-infected cell lysate.

**Fig. 1.**
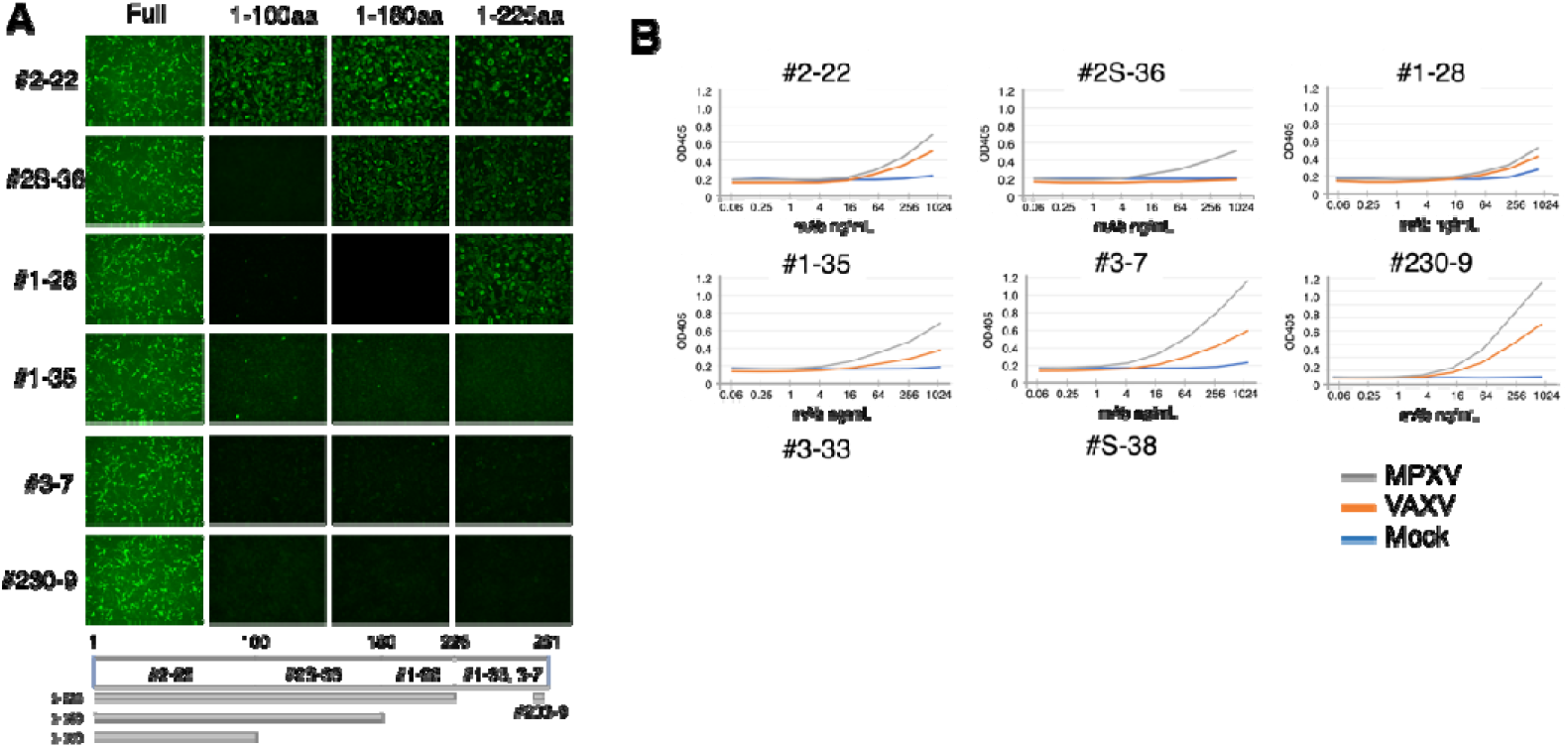
Antibody development for MPXV detection. (A) Immunofluorescence images of HEK293 cells expressing a series of C-terminal deletion mutants of MPXV A5L stained with the indicated mAbs. A schematic diagram of the deletion mutants of MPXV A5L; the locations of the epitopes for each mAb are shown. (B) ELISA screening of mAbs using MPXV (SPL-2A7, a clade IIb strain isolated in 2022 in Japan), VACV (strain LC16m8), or mock-infected RK13 cell lysates as the antigen. The cells were lysed in 1% NP40/PBS, and the supernatants, which were diluted 1000-fold in PBS, were added to each well of a 96-well plate. Serially diluted mAbs were added to the plates. The figure reports the values from a single experiment.

These results indicate that #2S-36 is a suitable antibody that can differentiate MPXV from VACV. An ELISA was performed to select clones to be paired with #2S-36. The results revealed that #1-28, #3-7, #1-35, and #230-9 had nearly identical sensitivities for MPXV A5L, whereas 2-22 did not (Supplemental Fig. 1A). Among these clones, #230-9, which recognizes the C-terminus of MPXV A5L and partially cross-reacted with VACV, was selected.

### 3.2 Development of a rapid antigen test for the detection of MPXV

The #2S-36/#230-9 set was tested in immunochromatography pairs, with one antibody conjugated to cellulose nanoparticles and one antibody absorbed into a nitrocellulose membrane. The test strips prepared by colloidal cellulose labeling with #230-9 and T□lines with #2S-36 presented greater detection sensitivity than the reverse combination did (Supplemental Fig. 1B). The kinetic analyses by surface plasmon resonance highlighted the fast association (kon) and slow dissociation (koff) rates for the interaction with MPXV A5L, resulting in equilibrium dissociation constants (K_D_ values) of 0.82 x10^−9^ M for #230-9 and 1.5 x10^−9^ M for #2S-36, respectively (Supplemental Fig. 2). Based on the test strips assay, an Ag-RDT was established with #2S-36 used as a capture Ab in the test (T) line and colloidal cellulose labeled #230-9 as a detection Ab absorbed on a pad. The control (C) line was incubated with a goat anti-rabbit IgG antibody. The analytical performance of the Ag-RDT was evaluated by testing the recombinant MPXV A5L as a standard sample in the concentration range of 0 to -1000 ng/mL. Naked-eye observation revealed that the A5L concentration of 0.5 ng/mL caused a slight but distinguishable difference in the test line intensity compared with that of the negative control (Fig. 2).

**Fig. 2.**
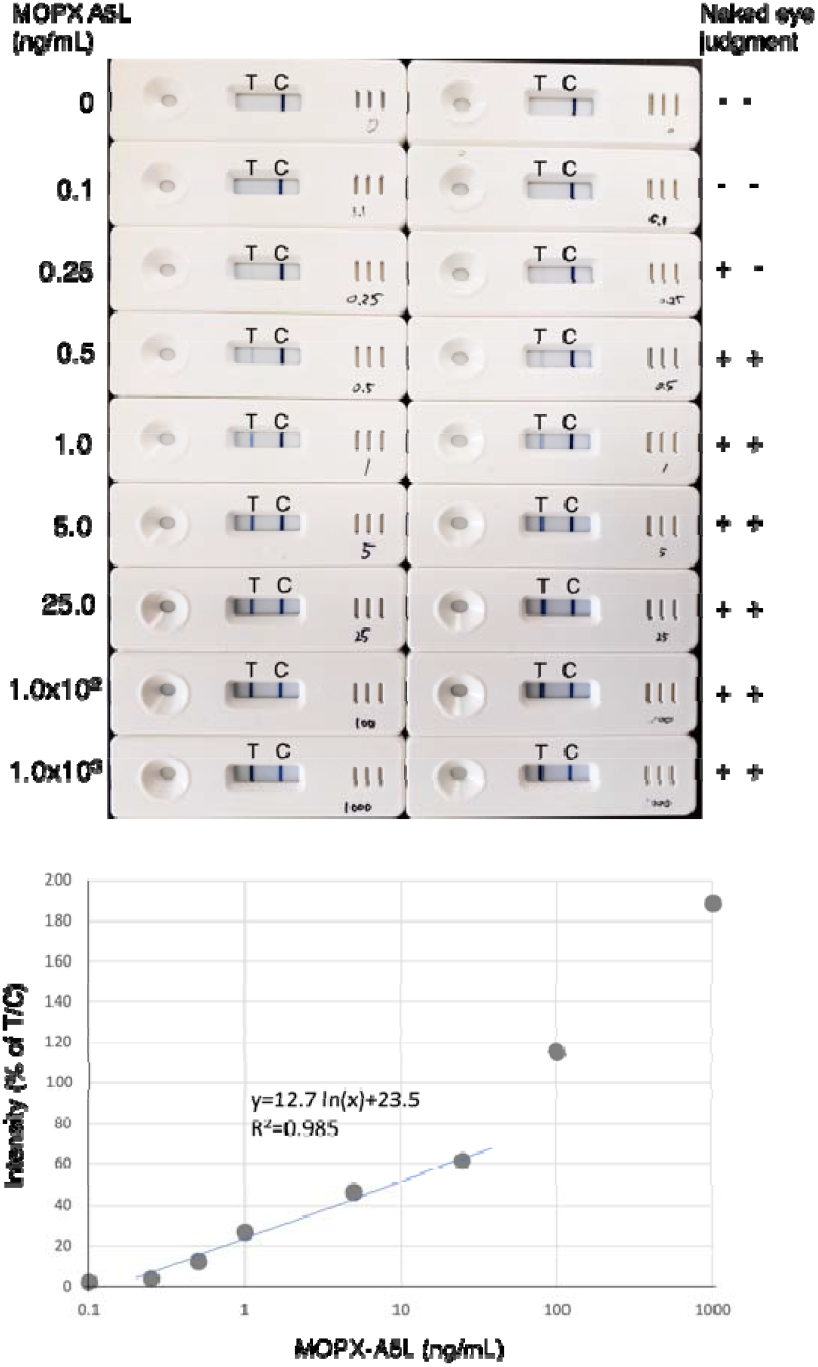
Limit of detection of the MPXV Ag-RDT using recombinant MPXV A5L as the antigen. (A) Photographs representing the LFIA strips and test zones. Recombinant MPXV A5L diluted with sample diluent solution and an aliquot of 110 μL was dropped into the immunochromatographic device. After 15 minutes, the presence of lines was visually checked. Immediately after visual inspection, images were captured, and the signal intensity at both the test and control line locations was quantified via ImageJ. The results of two experimental replicates are shown. The visual limit of detection was set by two independent observers at the lowest A5L concentration at which the T line disappeared in the two strips. Naked-eye judgment: positive (+) or negative (-). C, control. T, test. (B) Standard curve plot of the T/C value (%) on the Y-axis and the logarithm of MPXV A5L on the X-axis.

A standard curve was generated by plotting the relative band intensity ratio (T/C) against the MPXV A5L concentration, which increased with increasing protein concentration. It was quantitative in the range of 0 to 25.0 ng/mL, with a correlation coefficient of 0.9994. Increasing the antigen concentration above 1000 ng/mL led to a decrease in the intensity of the C line, whereas the intensity of the T line continued to increase, affecting the T/C ratio. This is likely due to fewer free-capture antibodies binding to the C line, as more antibodies are used to form immune complexes. The limit of detection (LOD) of the Ag-RDT was 440 plaque-forming units (PFU)/test for clade IIb SPL-2A7 and 2112 PFU/test for clade I Congo8 virus. Cross-reactivity with VACV was not detected at concentrations under 1.76×10^4^ PFU/test (Fig. 3). The Ag-RDT did not cross-react with viruses causing diseases with symptoms similar to those of MPXV, such as the measles virus, Coxsackie virus type A6, A16, and enterovirus A71 (Supplemental Fig. 3). The Ag-RDT was negative for all 20 bacterial lysates present in the airway and oral cavity (Supplemental Fig. 4).

**Fig. 3.**
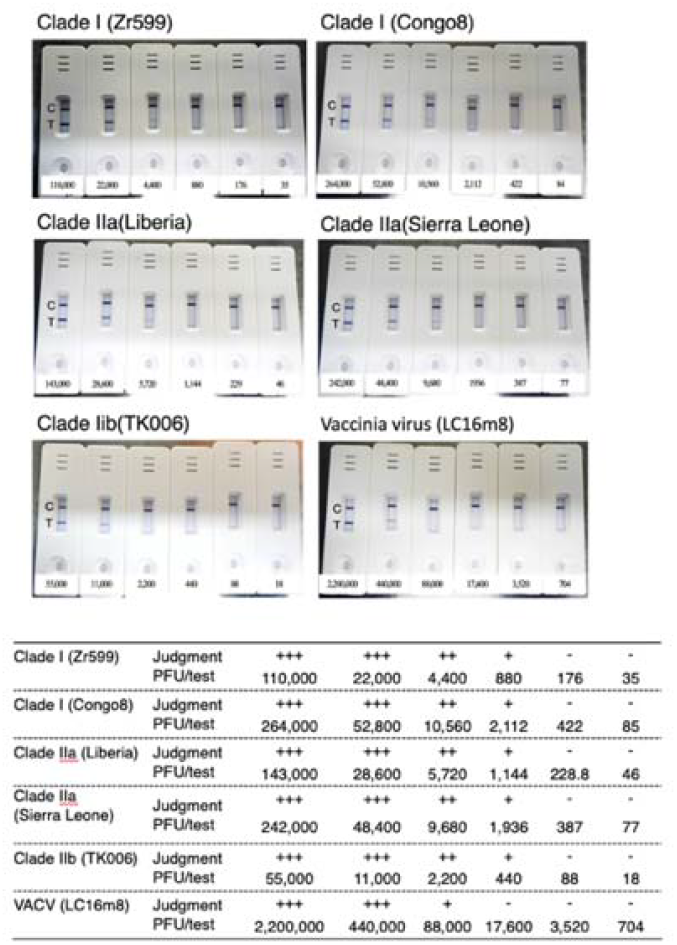
Analytical sensitivity and specificity of the MPXV Ag-RDT using serially diluted stocks of isolates of MPXV or VACV. An aliquot of 75 μl of the viral mixture was diluted with 75 μl of sample dilution buffer, and 110 μl was subsequently added to the cassette. Visual judgment was independently determined by two people.

When concentrations of human or bovine serum, ranging from 0.2% to 8%, were spiked with MPXV (11,000 PFU) and then applied to the Ag-RDT, no significant inhibition of the test line intensity was observed, indicating that the serum did not significantly affect the performance of the assay (Supplemental Fig. 5).

### 3.3 Point-of-care clinical evaluation studies

We next conducted a study to compare the sensitivity of the Ag-RDT with that of qPCR for MPXV clinical specimens collected as dry swabs (Kasuya et al., 2023). The qPCR-positive samples were divided into high (Ct value of ≤□25) and low (Ct value of 26∼40) groups. As shown in Fig. 4 and Table 1, The Ag-RDT detected 87% (20 out of 23 swabs; 95% CI, 66%-97%) of the samples in the high group but failed to detect samples in the low group (0 out of 27 swabs; 95% CI, 0%-13%). The Ag-RDT results were consistent with those of the qPCR-negative samples, with a specificity of 100% (0 out of 10 swabs; 95% CI, 0%-31%).

**Fig. 4.**
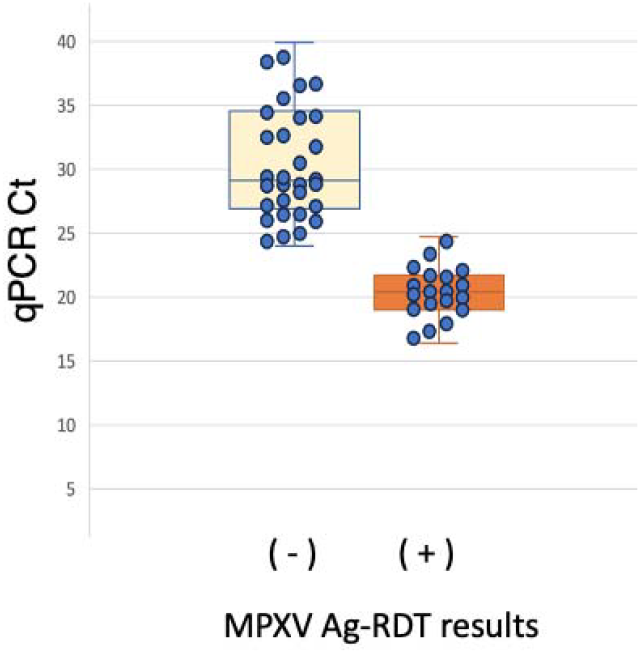
The MPXV Ag-RDT-positive and antigen-negative samples were compared via a box plot of the qPCR Ct values. Each symbol represents one sample. The center lines show the medians. The box limits are quartiles 1 and 3, and the whiskers represent the maximum and minimum values.

## 4. Discussion

In this study, we designed an MPXV Ag-RDT by developing an antibody set that can differentiate MPXV from VACV. The Ag-RDT provided accurate results in less than 15 minutes and exhibited high selectivity and analytical sensitivity for MPXV. A concordance study of clinical specimens collected as dry swabs revealed that the Ag-RDT showed 87% sensitivity in samples with Ct values of ≤□25 and a specificity of 100% with qPCR-negative samples.

Since the global outbreak of monkeypox in 2022, combined with the increasing danger of other zoonotic orthopoxvirus infections, there has been a need to develop a MPXV-specific diagnostic tests. Among the MPXV proteins, the virus envelope protein A29, which plays an important role in virus□host cell interactions, is one of the most extensively studied targets (Ahsendorf et al., 2019; Bengali, Satheshkumar, and Moss, 2012; Howard, Senkevich, and Moss, 2008). Several in-house and commercial Ag-RDTs have been developed by using antibodies specific to A29. However, most of these are cross-reactive with VACV (de Lima et al., 2023; Dubois, Hammarlund, and Slifka, 2012). Owing to the increasing incidence of occasional outbreaks of VACV in South America, the use of such tests can result in misdiagnosis as MPXV (Miranda et al., 2017). Recently, mAbs that can distinguish MPXV from VACV have been developed and applied to Ag-RDTs (Davis et al., 2023; Hughes et al., 2014; Ye et al., 2023). Although MPVX typically displays a low number of mutations, such as those associated with DNA viruses, the mutation ratio of MPVX isolated after the 2022 outbreak surpassed the standard mutation rate of the virus (Yashavarddhan et al., 2023). Consequently, antibodies that recognize the A29 epitope may not work as effectively in the future because of viral mutations, which could cause decreased sensitivity and inaccurate results. To prepare for unexpected mutant strains of MPXV that may evade current tests, it is crucial to develop an additional set of antibodies targeting a different protein of MPXV.

Among orthopoxviruses, MPXV A5L is one of the major core immunogenic and conserved proteins (Risco et al., 1999). The two antibodies targeting MPXV A5L are suitable for use in rapid diagnostic tests because they can bind to a distant epitope with a high association rate constant (kon) and a low dissociation rate constant (koff). These characteristics enable the antibodies to quickly bind to the antigen and maintain strong binding. This makes them ideal for designing species-specific serological assays targeting MPXV A5L. No cross-reactivity was observed for viruses causing diseases with symptoms similar to those of monkeypox, bacteria present in the airway and oral cavity and interfering substances in the serum. No loss of sensitivity was observed in this test when the samples were stored at room temperature for one year after manufacturing.

The viral load has been found to be the most important factor for determining antigen test sensitivity (Lim et al., 2023). Our data showed that the Ag-RDT has enough sensitivity to identify contagious patients with Ct ≤ 25. Therefore, this test could guide initial decisions regarding isolation and management and reduce the need for a qPCR test in the case of positive results. It is important to note that the sensitivity of the test decreased when the qPCR Ct value was greater than 25. As a result, this test may not be appropriate for detecting the virus in the prodromal stage, as the viral load may be too low to be detected by the Ag-RDT. Prior to clinical implementation, this device requires several improvements to enhance its sensitivity and performance.

## Conclusion

The MPXV Ag-RDT is capable of sensitive and specific detection of MPXV in virus-infected clinical specimens, distinguishing it from other skin infections. The MPXV Ag-RDT has potential for use in point-of-care settings.

## Data Availability

All data produced in the present study are available upon reasonable request to the authors.

## Abbreviations

Ag-RDT: antigen-detecting rapid diagnostic test
MPXV: monkeypox virus
OPXV: orthopoxvirus
qPCR: quantitative polymerase chain reaction
LFIA: lateral flow immunochromatography assay
VACV: vaccinia viruses
Ct: cycle threshold

## Transparency declaration

All the authors have nothing to disclose.

## CRediT authorship contribution statement

Nobuyuki Kurosawa: Writing – original draft, Methodology, Investigation, Formal analysis, Data curation. Tatsuhiko Ozawa: Methodology, Investigation, Formal analysis. Madoka Kawahara: Methodology, Investigation, Formal analysis. Masayuki Shimojima: Methodology, Investigation, Formal analysis. Madoka Kawahara: Methodology, Investigation, Formal analysis. Fumi Kasuya: Methodology, Investigation, Formal analysis. Wakaba Okada: Methodology, Investigation, Formal analysis. Mami Nagashima: Methodology, Investigation, Formal analysis. Kenji Sadamasu: Methodology, Investigation, Formal analysis. Masae Itamochi: Methodology, Investigation, Formal analysis. Hideki Tani: Methodology, Investigation, Formal analysis. Yoshitomo Morinaga: Methodology, Investigation, Formal analysis. Masaharu Isobe: Funding acquisition, Supervision.

## Data availability

The data will be made available upon request.

## Acknowledgments

We thank past and current members of our laboratory for fruitful discussions. We also thank Y. Nohara and K Takai for their technical support.

