## Supplemental file for "Development and clinical evaluation of a monkeypox antigen-detecting rapid diagnostic test"

Nobuyuki Kurosawa

Department of Life Sciences and Bioengineering, Laboratory of Molecular and Cellular Biology, Faculty of Engineering, Academic Assembly, University of Toyama, Toyama, Japan

**Abstract**

To address the global emergence of monkeypox after the 2022 epidemic, a rapid and accurate diagnostic tool is needed at the point of care to identify individuals infected with monkeypox virus (MPXV) to prevent and control the spread of the virus. We designed an antigen-detecting rapid diagnostic test that exclusively detects MPXV without cross-reacting with the vaccinia virus by developing monoclonal antibodies against the MPXV nuclear capsid protein A5L (MPXV-A5L). The test established a limit of detection sensitivity of 0.5 ng/mL of MPXV-A5L, with high sensitivity (87%) for clinical specimens collected from MPXV patients, a qPCR cycle threshold value ≤ 25 and 100% specificity for qPCR-negative samples. The test is an ideal rapid diagnostic tool for supporting clinical decision-making for people suspected of having MPXV infection in resource-poor settings.

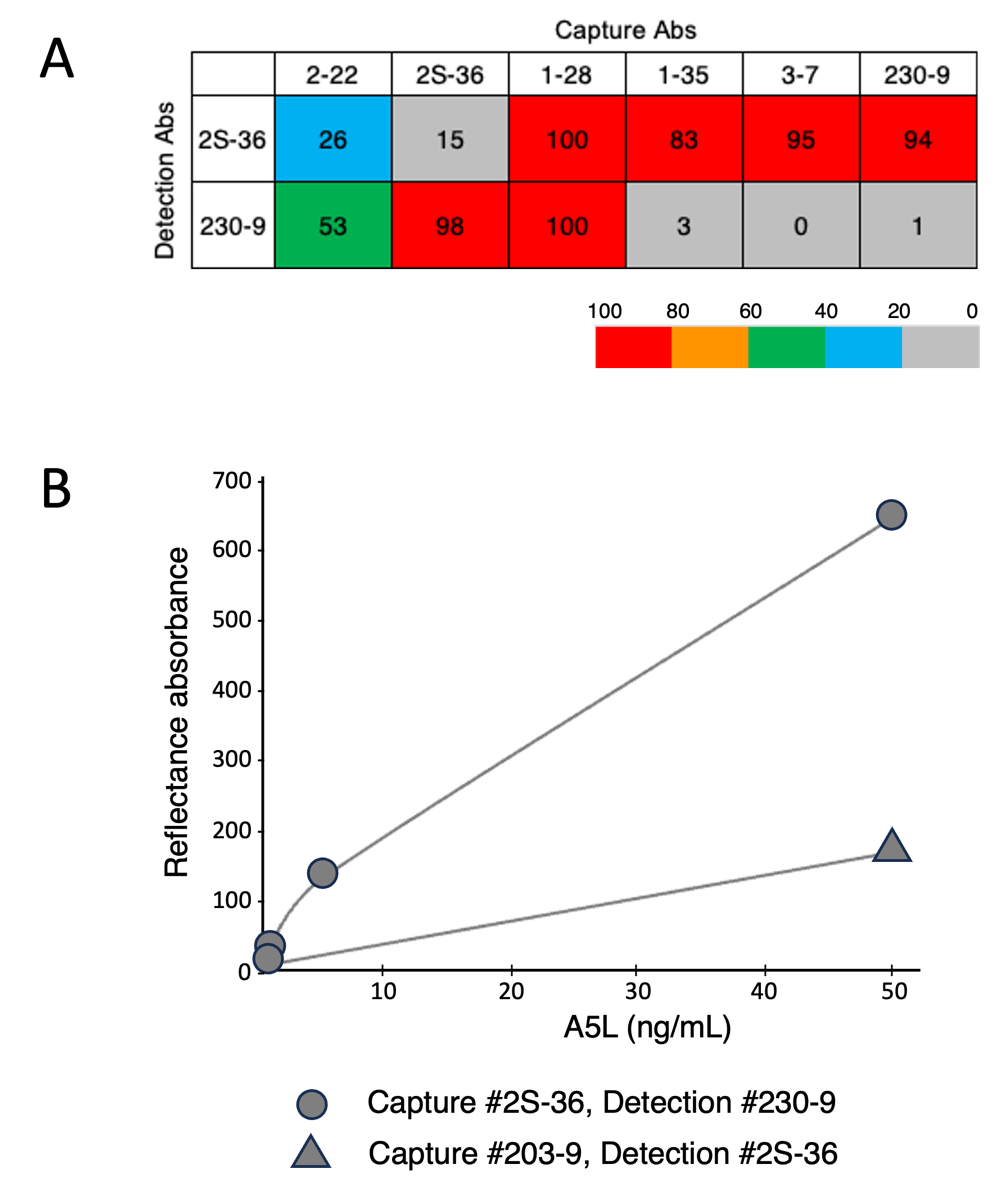

Supplemental Fig.1 (A) Pairwise coupling of six mAbs against #2S-36 or #230-9. Each well of 96-well plates immobilized with a capture antibody (10 ng) was incubated with 10 ng of MOPX-A5L for 1 h. After washing the plate with PBST, a detection antibody (10 ng/mL) labeled with alkaline phosphatase was added to each well and incubated for 30 min. The signal was developed with Bluephos and read at OD_600_ nm. The performance of the antibody pair is shown in the heatmap. The numbers inside the grid represent the relative performance of the antibody pair. Data are expressed as the mean of three measurements. (B) The test strips prepared by colloidal cellulose labeling with #230-9 and T‐lines with #2S-36 were compared for detection sensitivity compared to the reverse combination.

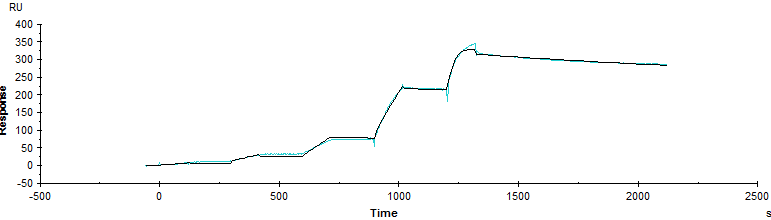

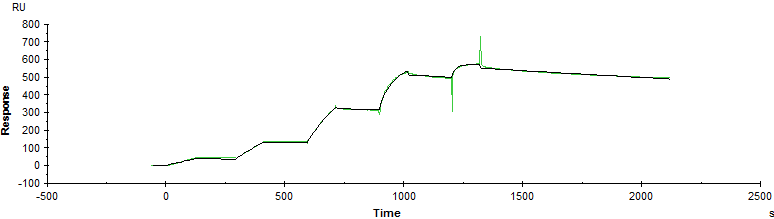

K_on_ = 9.63 x 10^5^ M^-1^ S^-1^

K_off_ = 1.49 x 10^-4^ S^-1^

K_D_ = 1.5 x10^–9^ M

K_on_ = 1.79 x 10^5^ M^-1^ S^-1^

K_off_ = 1.48 x 10^-4^ S^-1^

K_D_ = 0.83 x10^–9^ M

#230-9

#2S-36

Supplemental Fig.2 Affinity measurement of #2S-36 and #230-9. Association and dissociation of each mAb to recombinant MPOX-A5L at various concentrations (50,25,12.5,6.25,3.13,1.56, and 0.78nM) were evaluated using Biacore 100 (two technical replicates).

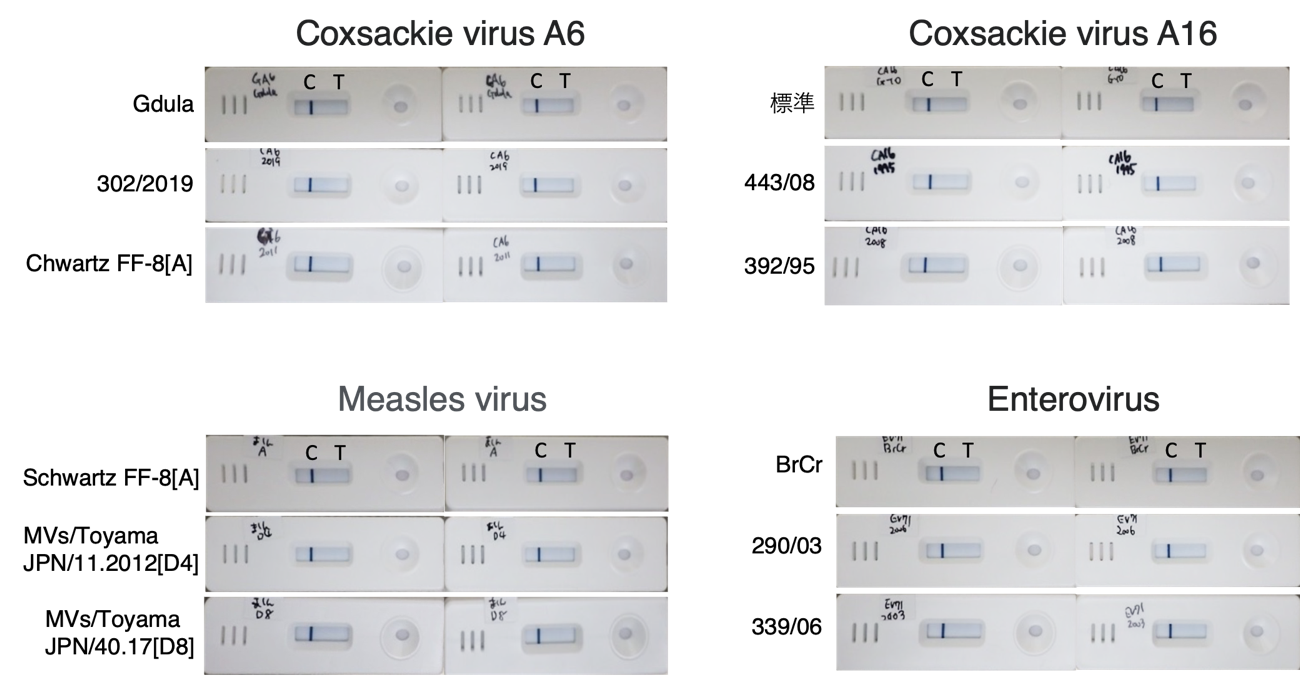

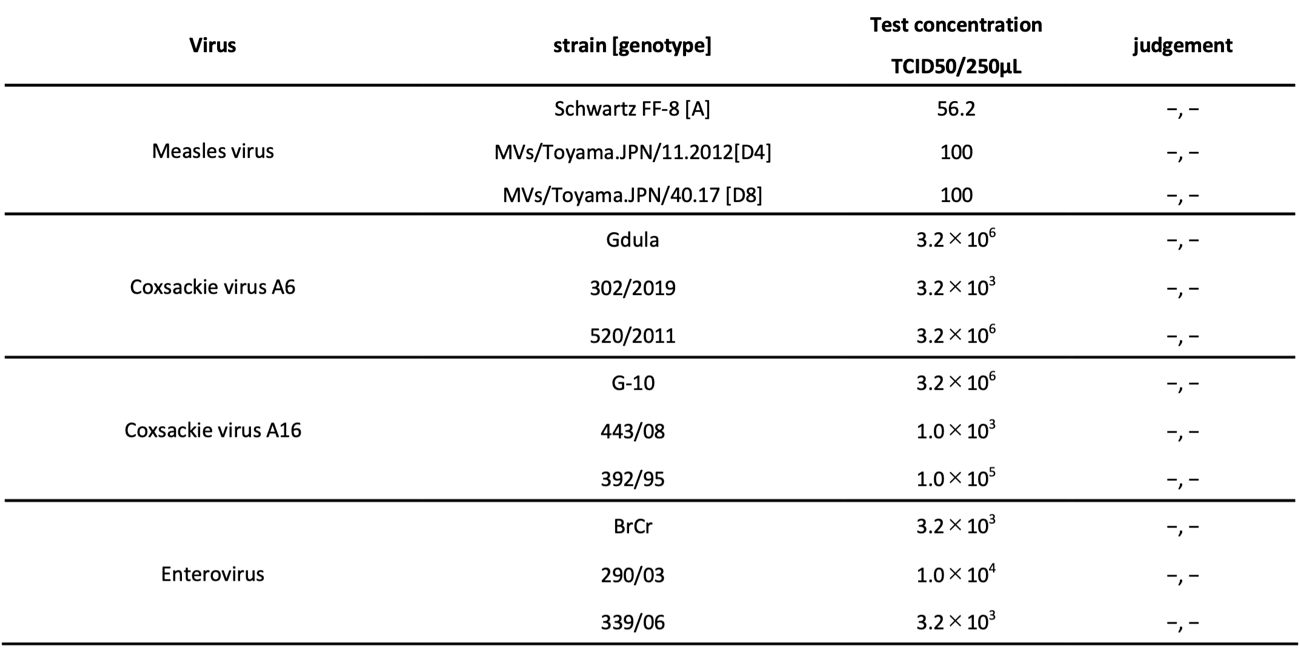

Supplemental Fig.3 MPXV Ag-RDT did not cross-react with viruses causing diseases with similar symptoms to MPOX viruses. An aliquot of 110 μL of the indicated virus solution was added to the kit.

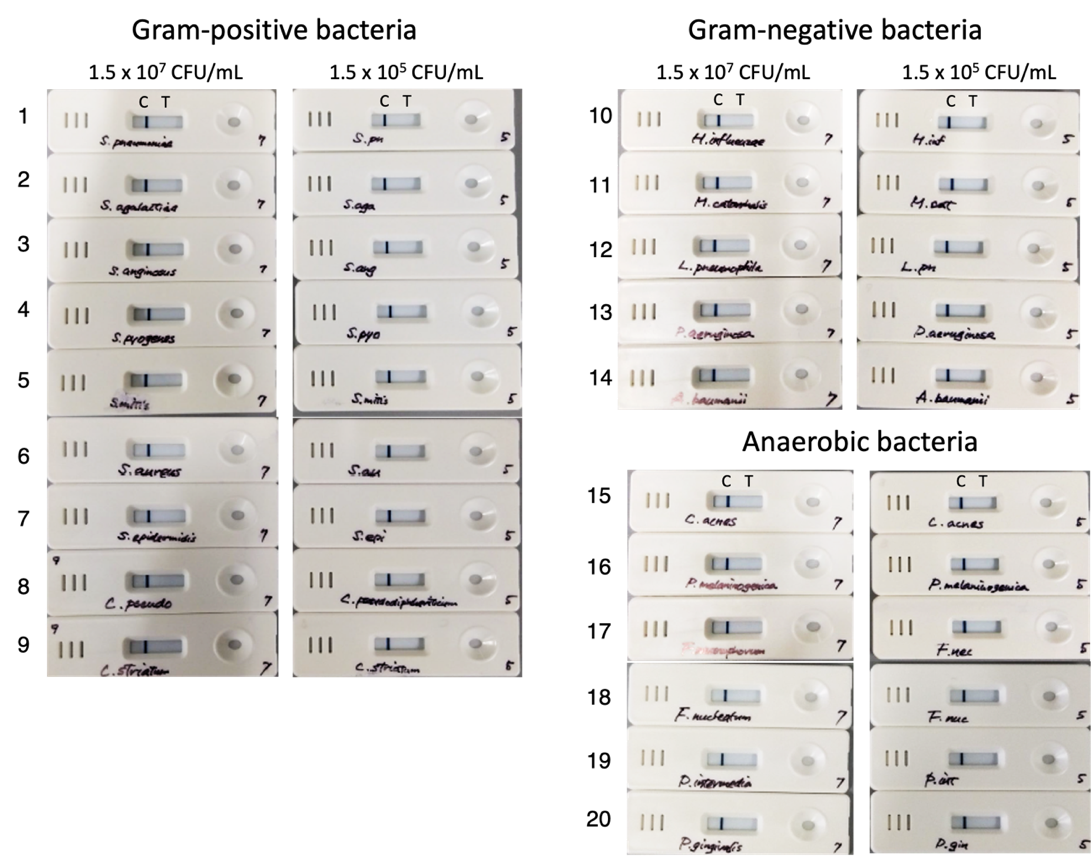

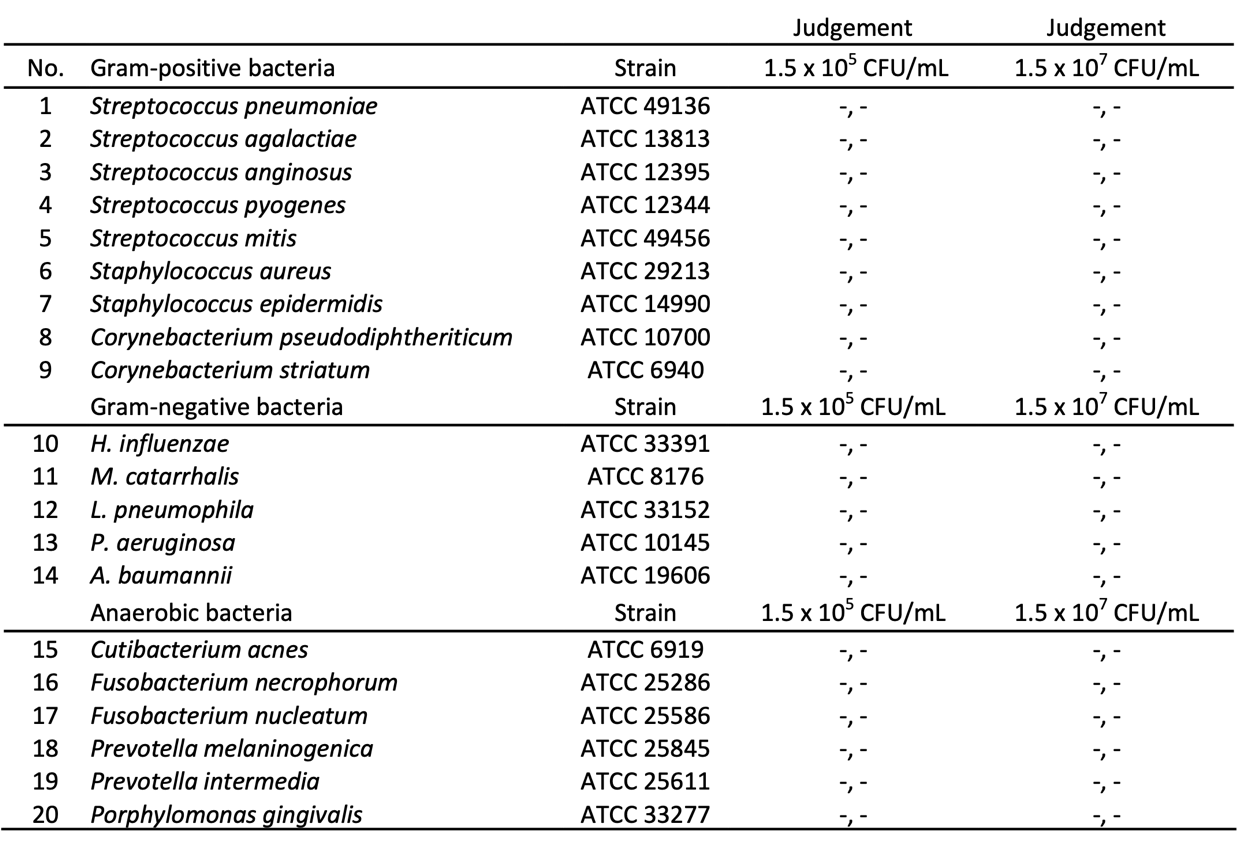

Supplemental Fig.4 MPXV Ag-RDT did not cross-react with bacteria present in the airway and oral cavity. Bacterial colonies on the plate were suspended in saline to adjust to a bacterial concentration of McF 0.5 ± 0.15 (equivalent to 1.5x10^8^ CFU/mL). This was diluted with sample dilution solution to adjust a bacterial concentration of 1.5x10^7^ colony-forming units per mL (CFU/mL) or 1.5x10^5^ CFU/mL, and 110 μL was added to the kit.

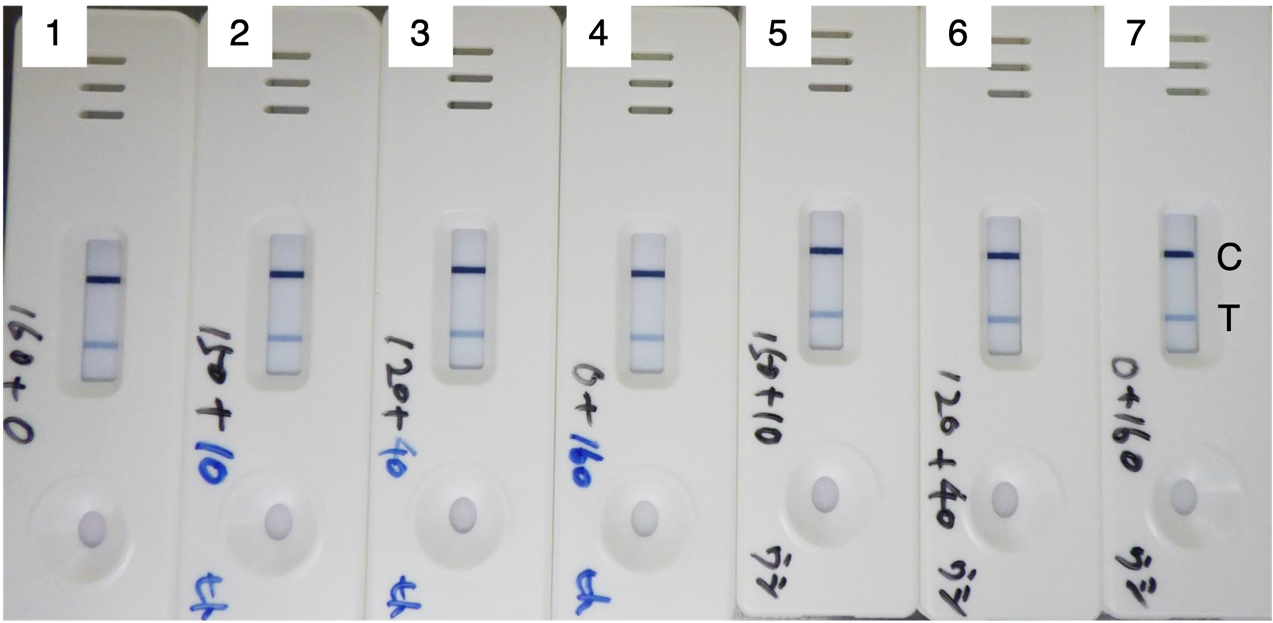

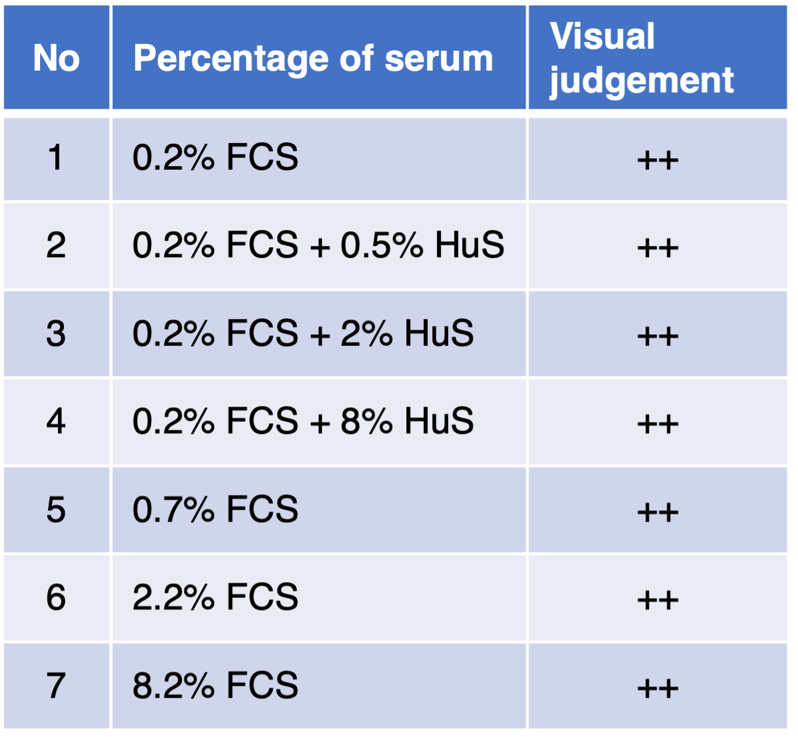

Supplemental Fig.5 Specificity of MPXV Ag-RDT for negative serum. Analytical performance of MPXV-LFIA using 11,000 PFU MPXV spiked into the indicated concentration of serum are shown.

Table 1. Clinical performance evaluation of MPXV Ag-RDT in comparison to qPCR. Clinical specimens were mixed with 0.5 mL of PBS. The aliquot of 75㎕ was diluted with 75㎕ of sample dilution buffer and then 110㎕ was applied on the cassettes.

| Sample No | qPCR Ct | MPXV Ag-RDT |  | Sample No | qPCR Ct | MPXV Ag-RDT |
| --- | --- | --- | --- | --- | --- | --- |
| 162 | 16.4 | ( + ) |  | 288 | 24 | ( - ) |
| 258 | 17 | ( + ) |  | 37 | 24.5 | ( - ) |
| 206 | 17.9 | ( + ) |  | 122 | 24.7 | ( - ) |
| 260 | 18.9 | ( + ) |  | 130 | 25.9 | ( - ) |
| 131 | 19 | ( + ) |  | 295 | 26 | ( - ) |
| 29 | 19.4 | ( + ) |  | 250 | 26.3 | ( - ) |
| 176 | 16.9 | ( + ) |  | 60 | 26.4 | ( - ) |
| 112 | 21 | ( + ) |  | 102 | 27.1 | ( - ) |
| 19 | 20.3 | ( + ) |  | 161 | 27.3 | ( - ) |
| 85 | 20.4 | ( + ) |  | 36 | 27.6 | ( - ) |
| 100 | 20.5 | ( + ) |  | 111 | 28.6 | ( - ) |
| 87 | 20.8 | ( + ) |  | 164 | 28.9 | ( - ) |
| 144 | 21 | ( + ) |  | 171 | 28.9 | ( - ) |
| 204 | 21.6 | ( + ) |  | 217 | 29 | ( - ) |
| 223 | 21.6 | ( + ) |  | 262 | 29 | ( - ) |
| 155 | 21.7 | ( + ) |  | 110 | 29.2 | ( - ) |
| 267 | 22.2 | ( + ) |  | 305 | 29.2 | ( - ) |
| 17 | 22.5 | ( + ) |  | 118 | 29.6 | ( - ) |
| 255 | 23.5 | ( + ) |  | 184 | 30.7 | ( - ) |
| 82 | 24.5 | ( + ) |  | 145 | 31.9 | ( - ) |
|  |  |  |  | 21 | 33 | ( - ) |
| * : VZV positive, Ct=18.3 | |  |  | 106 | 33 | ( - ) |
| ND : not detected | |  |  | 287 | 34.5 | ( - ) |
| ( + ) : Positive | |  |  | 154 | 34.6 | ( - ) |
| ( - ) : Negative | |  |  | 241 | 34.9 | ( - ) |
|  |  |  |  | 167 | 36.2 | ( - ) |
|  |  |  |  | 50 | 37.4 | ( - ) |
|  |  |  |  | 183 | 37.6 | ( - ) |
|  |  |  |  | 153 | 39.4 | ( - ) |
|  |  |  |  | 24 | 39.9 | ( - ) |
|  |  |  |  | 165* | ND | ( - ) |
|  |  |  |  | 28 | ND | ( - ) |
|  |  |  |  | 86 | ND | ( - ) |
|  |  |  |  | 107 | ND | ( - ) |
|  |  |  |  | 139 | ND | ( - ) |
|  |  |  |  | 170 | ND | ( - ) |
|  |  |  |  | 190 | ND | ( - ) |
|  |  |  |  | 211 | ND | ( - ) |
|  |  |  |  | 243 | ND | ( - ) |
|  |  |  |  | 296 | ND | ( - ) |
